# Reducing diagnostic delays in Acute Hepatic Porphyria using electronic health records data and machine learning: a multicenter development and validation study

**DOI:** 10.1101/2023.08.30.23293130

**Authors:** Balu Bhasuran, Katharina Schmolly, Yuvraaj Kapoor, Nanditha Lakshmi Jayakumar, Raymond Doan, Jigar Amin, Stephen Meninger, Nathan Cheng, Robert Deering, Karl Anderson, Simon W. Beaven, Bruce Wang, Vivek A. Rudrapatna

## Abstract

**Importance:** Acute Hepatic Porphyria (AHP) is a group of rare but treatable conditions associated with diagnostic delays of fifteen years on average. The advent of electronic health records (EHR) data and machine learning (ML) may improve the timely recognition of rare diseases like AHP. However, prediction models can be difficult to train given the limited case numbers, unstructured EHR data, and selection biases intrinsic to healthcare delivery.

**Objective:** To train and characterize models for identifying patients with AHP.

**Design, Setting, and Participants:** This diagnostic study used structured and notes-based EHR data from two centers at the University of California, UCSF (2012-2022) and UCLA (2019-2022). The data were split into two cohorts (referral, diagnosis) and used to develop models that predict: 1) who will be referred for testing of acute porphyria, amongst those who presented with abdominal pain (a cardinal symptom of AHP), and 2) who will test positive, amongst those referred. The referral cohort consisted of 747 patients referred for testing and 99,849 contemporaneous patients who were not. The diagnosis cohort consisted of 72 confirmed AHP cases and 347 patients who tested negative. Cases were female predominant and 6-75 years old at the time of diagnosis.

Candidate models used a range of architectures. Feature selection was semi-automated and incorporated publicly available data from knowledge graphs.

**Main Outcomes and Measures:** F-score on an outcome-stratified test set

**Results:** The best center-specific referral models achieved an F-score of 86-91%. The best diagnosis model achieved an F-score of 92%. To further test our model, we contacted 372 current patients who lack an AHP diagnosis but were predicted by our models as potentially having it (≥ 10% probability of referral, ≥ 50% of testing positive). However, we were only able to recruit 10 of these patients for biochemical testing, all of whom were negative. Nonetheless, *post hoc* evaluations suggested that these models could identify 71% of cases earlier than their diagnosis date, saving 1.2 years.

**Conclusions and Relevance:** ML can reduce diagnostic delays in AHP and other rare diseases. Robust recruitment strategies and multicenter coordination will be needed to validate these models before they can be deployed.

**KEY POINTS:** *Question:* Can machine learning help identify undiagnosed patients with Acute Hepatic Porphyria (AHP), a group of rare diseases?

*Findings:* Using electronic health records (EHR) data from two centers we developed models to predict: 1) who will be referred for AHP testing, and 2) who will test positive. The best models achieved 89-93% accuracy on the test set. These models appeared capable of recognizing 71% of the cases earlier than their true diagnosis date, reducing diagnostic delays by an average of 1.2 years.

*Meaning:* Machine learning models trained using EHR data can help reduce diagnostic delays in rare diseases like AHP.

## INTRODUCTION

The paradox of rare diseases is that they collectively affect as much as 10% of the population yet are individually uncommon, with a prevalence under 1:2000^1^. Diagnosing them requires significant expertise, which is typically a limiting factor. Patients with rare diseases can suffer for years or decades without a diagnosis, leading to unnecessary tests, ineffective (or even harmful) treatments, lost quality of life, underemployment, and other costs affecting individuals and society.

An example of this is Acute Hepatic Porphyria (AHP), a group of rare but treatable, inherited diseases of heme biosynthesis. AHP presents with episodes of severe pain that result from the abnormal accumulation of delta-aminolevulinic acid (d-ALA) and porphobilinogen (PBG)^2^. AHP attacks are episodes of neurologic damage, and patients with repeated attacks are at risk for complications affecting the nervous system, liver, and kidneys^3–5^. The prevalence of symptomatic AHP is roughly 1:100,000^6,7^, contributing to average diagnostic delays of 15 years following symptom onset ^8^. Thus, many AHP patients have high chronic disease burden and poor quality of life^9,10^. However, the FDA approval of givosiran now provides a prophylactic option for AHP patients with recurrent attacks. An important next step is to reduce diagnostic delays in these patients so they may manage AHP earlier and potentially prevent downstream morbidity and even mortality^9^.

Advances in machine learning (ML) and methods for using electronic health records (EHR) data offer an alternative to the standard of care, which relies on local expertise to make these diagnoses. EHRs contain detailed information on the patient journey, including symptoms, (mis)diagnoses, and attempted treatments prior to a successful diagnosis. These data could help train ML models to identify these patients earlier^11^.

Despite this promise, building accurate and generalizable models for patient identification remains difficult. The low prevalence of these patients results in class imbalance, where models have too few cases and too many controls for optimal learning, particularly in the face of many predictive features. Moreover, EHR data accumulates over time as patients navigate through health systems in ways that influence their chances of being correctly diagnosed. Thus, a careful attention to time (to prevent models from using future information to predict the past) and to possible selection biases along the patient journey is critical to avoid errors that have affected prior efforts^12–14^.

Here we developed an ML approach to reduce diagnostic delays in AHP. Using the EHR data from two medical centers (UCSF, UCLA), we trained models to sequentially predict the likelihood that a patient would eventually be tested for AHP, and the likelihood that they would test positive. We evaluated the models on a hold-out test set and characterized their potential impact on the patient journey. Lastly, we attempted a prospective validation of these models by recruiting and biochemically testing current patients who were predicted as having AHP.

## METHODS

The study was approved by the institutional review boards at UCSF (20-31754) and UCLA (21-001260).

### Cohort selection

We constructed a causal model of the patient journey towards a new diagnosis (Figure 1). It revealed that this journey could be subdivided into two key “selection” steps: 1) the decision to refer a patient for AHP testing, and 2) confirmation of a diagnosis. Importantly, these steps were associated with predictors that were likely to be captured within EHR systems. However, the predictors were thought to be different for each step. For example, predictors of a clinician’s likelihood of considering an AHP diagnosis and/or of patients encountering such a clinician may be different from the predictors that dictate the AHP diagnosis itself. We decided to model each step using different cohorts, one subject to the decision to refer (“referral cohort”), and the other with a chance of testing positive (“diagnosis cohort”).

**Figure 1:**
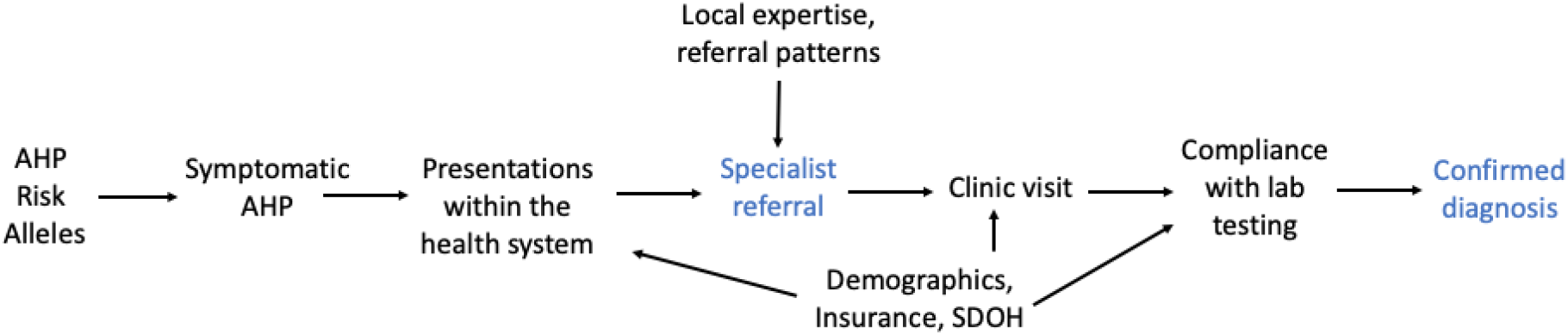
A causal model of the AHP patient journey to a confirmed diagnosis. The probability of a within-system new diagnosis of AHP is influenced by multiple factors, including the number and nature of patient presentations, local expertise of providers, availability of specialists and/or lab testing, and patient’s ability and desire to follow clinical recommendations. We identified two selection “bottlenecks”: the probability that a patient will be identified by a given provider as needing a specialty evaluation for possible AHP, and the probability that a tested patient will be confirmed as a new case (blue font). Each of these bottlenecks were separately modeled, to reduce bias and improve the accuracy of downstream predictions.

**Figure 2:**
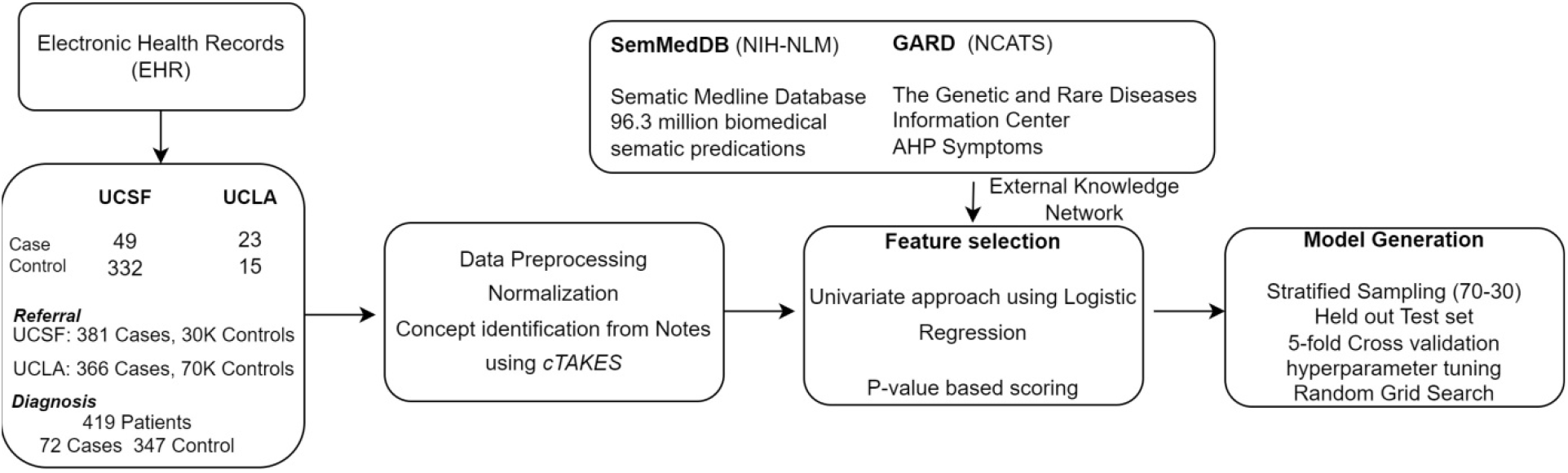
An overview of the ML pipeline. EHR data from UCSF and UCLA were harmonized to generate Referral and Diagnosis cohorts. The structured data from these patients underwent preprocessing and normalization, and the unstructured data were partially structured using cTAKES and regular expressions. These data were normalized and harmonized. We used data-driven and KG-based feature selection techniques to reduce the feature space from 25,320 candidate features to 138 final features. These were used to train, perform model selection using cross validation, and finalize models to be used for prospective validation.

We queried structured EHR databases (UCSF, 2012-2022; UCLA, 2019-2022) to identify cases and controls for each cohort. Referral cases were identified based on patients aged 10-65 with an encounter in the porphyria clinic (UCSF) or a completed AHP test (UCLA). Referral controls were patients with at least one clinical encounter for abdominal pain. This is the most common symptom of AHP attacks, and patients meeting this definition were conceptualized as having a non-zero probability of being tested.

Diagnosis cases were patients with AHP. These cases came from a disease registry (UCSF) and following manual review of patient records (UCLA). All cases had urinary d-ALA or PBG levels greater than twice the laboratory upper limit of normal, with compatible symptoms at the time of testing. At UCLA, potential cases subject to manual review were identified from the EHR database based on having one of the following elements: an AHP ICD code, a hemin order, or a urine d-ALA or PBG lab order. Cases were all confirmed by clinician experts on the study team. Diagnosis controls were patients who were either seen in the porphyria clinic (UCSF) or completed adequate biochemical testing at the time of compatible symptoms (UCLA), and were confirmed as a non-case.

### Data preparation

We harmonized EHR data from both centers to facilitate downstream modeling. These data consisted of standard structured data (diagnosis, procedures, medications, laboratory tests, demographics, encounters, providers) and machine-redacted notes^15^. We used cTAKES^16^ to extract named entities to serve as potential predictive features. We also used domain knowledge and regular expressions to generate additional text-based features. We preprocessed the structured data to generate one-hot-encoded features (Supplemental Methods).

We generated two datasets corresponding to the referral and diagnosis cohorts as described above. We then subject the data to a time-based filter to remove all future elements that would not be available to a deployed model at inference time. For the referral model, we removed all data elements that occurred on or after the date of referral to the UCSF porphyria clinic, or occurring on or after the date that a UCLA patient had a structured data element of interest (ICD code, hemin order, urine d-ALA/PBG lab order). Of note, data from the date of referral was also excluded to prevent models from predicting referrals using the information in concurrently documented “assessment and plan” sections, which would not be useful in practice. For the diagnosis model, we removed all data elements occurring on or after the date of the first biochemical evaluation of AHP. This was done to prevent information leakage (e.g. documentation indicating that the diagnosis was already known to the patient or clinician).

### Feature selection

The initial feature space included standard structured data and one-hot-encoded features derived from notes. The latter were generated using automated methods (Supplemental Methods) as well as manually, using domain knowledge. For each dataset (referral, diagnosis), we selected only those features meeting these criteria: 1) a Wald p-value < 0.05 on a single logistic regression model, and 2) features directly linked to AHP in knowledge graphs. We used two knowledge graphs (KGs): SemMedDB^17^, a database of information sourced from PubMed abstracts, and a custom knowledge graph generated from the Genetic and Rare Diseases Information Center^1^. The latter was generated using Graphvite^18^, which links entities to AHP if they are described as being associated with it. EHR-based features were retained if they matched a relevant KG-based entity based on shared UMLS concepts^19^.

Following this approach, we began with 25,320 candidate features, applied p-value based selection to reduce to 3,316 features, and used KGs to finalize 138 features.

### Model training, internal validation

We used each dataset (referral, diagnosis) to define a different prediction task: 1) who will be referred for testing, among those with some chance of being referred (i.e. an abdominal pain encounter), and 2) who will test positive, among those who underwent testing. We used scikit-learn^20^ and autoGluon^21^ to train and evaluate 87 model architectures (Supplemental Methods). Each dataset underwent outcome-stratified splitting, with 70% used for training and 5-fold cross-validation, and 30% for testing.

For referral prediction, we compared results using a combined dataset as well as using separate referral models from each center. We only generated one diagnosis model given the paucity of AHP cases. The models achieving the highest F1 score on the test set were retrained on the full dataset prior to downstream tasks.

### Prospective evaluation

We applied the referral and diagnosis models to recently seen patients at UCSF or UCLA (≥1 encounter in 2019-2022) meeting the following additional criteria: 1) aged 18-65, 2) opted into research, 3) having a patient-provider messaging account. We identified all patients with ≥ 10% referral probability and ≥ 50% diagnosis probability. These thresholds were selected to account for the low probability of referrals for rare conditions. We calculated the expected number of new diagnoses, under the assumption of a 20% response rate, to confirm non-futility of a potential prospective evaluation. We then utilized patient recruitment services at UCSF and UCLA to solicit patient participation in a prospective test of model accuracy. Patients were solicited via electronic messaging and mail. Consented participants submitted a urine specimen for d-ALA and PBG testing.

### Model characterization

We applied the finalized models to data from the UCSF AHP cases and evaluated if they could have identified any patients earlier than their actual diagnosis year, established by manual review. We retrospectively applied the model to each year of EHR data (2012-2022) and used data available at the end of that year to calculate the running probability of a positive diagnosis. We counted as a model diagnosis any patient meeting the same probability thresholds as the prospective study. We tabulated cases that were model-identifiable ≥ 1 years prior to their actual diagnosis, and quantified the mean years saved over the full cohort.

## RESULTS

We defined two cohorts to train sequential models that estimate 1) the probability that a patient will be referred for AHP testing, among those with a reasonable chance of being referred (“referral cohort”), and 2) the probability that a patient will test positive for AHP, among those who underwent definitive testing (“diagnosis cohort”). The referral cohort, corresponding to all patients with at least one clinical encounter for abdominal pain (a common symptom of AHP), consisted of 100,596 patients. 747 (381 from UCSF; 366 from UCLA) were “referral cases”, that is, patients who were referred for testing. The remainder were “referral controls” (29,963 at UCSF; 69,886 at UCLA). The diagnosis cohort consisted of 419 patients, with 72 confirmed AHP cases (49 from UCSF, 23 from UCLA), and 347 controls (332 from UCSF, 15 from UCLA).

Of note, the number of referral cases (747) was larger than the size of the diagnosis cohort (419) because many patients who were referred for testing did not complete it. In this analysis, we assumed that the population of patients who complied with this clinical recommendation for testing was a representative subset of the overall patient pool referred for testing. In other words, patients with an underlying (but probably unknown) diagnosis of AHP were neither more nor less likely to complete a confirmatory diagnostic test. This assumption was thought to be reasonable on *a priori* grounds, and enabled a more straightforward application of these models at inference time.

Our AHP cohort was enriched for non-Hispanic white females, with most diagnosed with AIP in their 20s or 30s. The sub-cohorts from UCSF and UCLA showed minor differences in clinical features (Table 1). For example, 29% at UCSF had a positive family history of AHP, compared to 13% at UCLA. These differences were expected given that UCSF is a tertiary-care center that receives many outside referrals. By contrast, UCLA has a broader primary care base, and thus more within system diagnoses of AHP.

**Table 1:** Demographics and clinical characteristics of the cases and controls from the diagnosis cohort. Cases correspond to patients with AHP. Controls correspond to patients who were tested and were deemed definitively negative for AHP. Percentages were rounded to the nearest whole number except in cases where the proportion was near 0.

| Characteristics | Case |  |  | Control |  |  |
| --- | --- | --- | --- | --- | --- | --- |
|  | Total<br>(N=72) | UCSF<br>Case<br>(N=49) | UCLA<br>Case<br>(N=23) | Total<br>(N=347) | UCSF<br>Control<br>(N=332) | UCLA<br>Control<br>(N=15) |
| Age 0-20 | 5 | 5 | 0 | 58 | 58 | 0 |
| Age 20-40 | 34 | 22 | 12 | 113 | 108 | 5 |
| Age 40-60 | 17 | 11 | 6 | 112 | 106 | 6 |
| Age 60+ | 16 | 11 | 5 | 64 | 60 | 4 |
| Age Min | 6 | 6 | 22 | 1 | 1 | 22 |
| Age Max | 74 | 74 | 73 | 88 | 88 | 77 |
| Female sex no.<br>(%) | 81 | 78 | 87 | 48 | 46 | 80 |
| Body-mass<br>index | 25.7±3.7 | 23.9±1.2 | 27.4±6.3 | 28.6±12.6 | 29.2±18.0 | 28.1±7.3 |
| Hispanic<br>Ethnicity | 6.9 | 8.1 | 6.2 | 15.27 | 14.15 | 4.1 |
| Non-Hispanic<br>Race |  |  |  |  |  |  |
| --White | 63 | 67 | 52 | 54 | 53 | 80 |
| --Black | 0.4 | 0.2 | 0.1 | 0.3 | 0.1 | 0.1 |
| --Asian | 0.1 | 0.8 | 0 | 0.2 | 0.2 | 0 |
| --Other | 38 | 32 | 48 | 46 | 47 | 20 |
| % with acute<br>recurrent<br>abdominal pain | 47 | 41 | 61 | 30 | 28 | 60 |
| % with a<br>positive family<br>history | 39 | 35 | 48 | 5 | 4 | 27 |
| % with a<br>psychiatric<br>diagnosis | 114 | 8 | 28 | 6 | 5 | 20 |
| % with Acute<br>intermittent<br>porphyria | 83 | 88 | 74 | - | - | - |
| % with<br>Hereditary<br>coproporphyrria | 10 | 8 | 13 | - | - | - |
| % with<br>Variegate<br>Porphyria | 7 | 4 | 13 | - | - | - |

We created separate referral machine learning models for each center, to account for differences in referral patterns by allowing for center-specific feature weights. We created a single diagnosis model to reduce overfitting and maximize predictive power.

### Model performance

For each task, we trained multiple ML models and evaluated them on an outcome-stratified test set. We selected the models achieving the highest F-score for downstream analysis. The highest performing models achieved an F-Score of 86% and 91% for predicting referrals, and an F-score of 92% (Table 2). An analysis of the most important features for the diagnosis model revealed a constellation of terms with a plausible connection to AHP (eFigure 1).

**Table 2:** Performance of the best machine learning models for the referral and diagnosis tasks. All numbers correspond to percentages rounded to the nearest whole number.

| Task | Model | F-Score | Accuracy | Sensitivity | Specificity | PPV |
| --- | --- | --- | --- | --- | --- | --- |
| UCSF Referral | Ada Boost | 91 | 94 | 92 | 85 | 84 |
| UCLA Referral | Logistic Regression | 86 | 89 | 91 | 79 | 86 |
| Diagnosis Model | Logistic Regression | 92 | 93 | 96 | 90 | 92 |

### Prospective evaluation

We used the finalized model architectures to predict new cases from the pool of undiagnosed patients (referral control group) that had recently been seen at UCSF or UCLA (2019-2022). We identified 406 patients with a referral probability of ≥ 10% and a diagnosis probability ≥ 50% (eFigure 2). We selected the 10% cutoff heuristically to balance two goals: 1) maximizing the power of a potential study to reject a null hypothesis corresponding to an unpredictive model, i.e. one that predicts all patients as having a 7/10,000 chance of AHP (the empiric probability in referral cohort), and 2) limiting model extrapolation over patients with essentially no chance of ever being tested for this condition. Within this group, and assuming a 20% probability of participation, we expected to identify 14 new cases (95% CI 7-24). As such, we decided to proceed with this prospective study.

After engaging subject recruitment services, we identified 372 patients meeting the following additional criteria: 1) alive and aged 18-65 as of the start of recruitment, 2) opted into research, 3) accessible via the myChart patient-provider portal (Figure 3). Following several rounds of recruitment, we consented 12 patients. 10 patients completed a confirmatory biochemical test, corresponding to a 2.7% participation rate. All patients tested negative.

**Figure 3:**
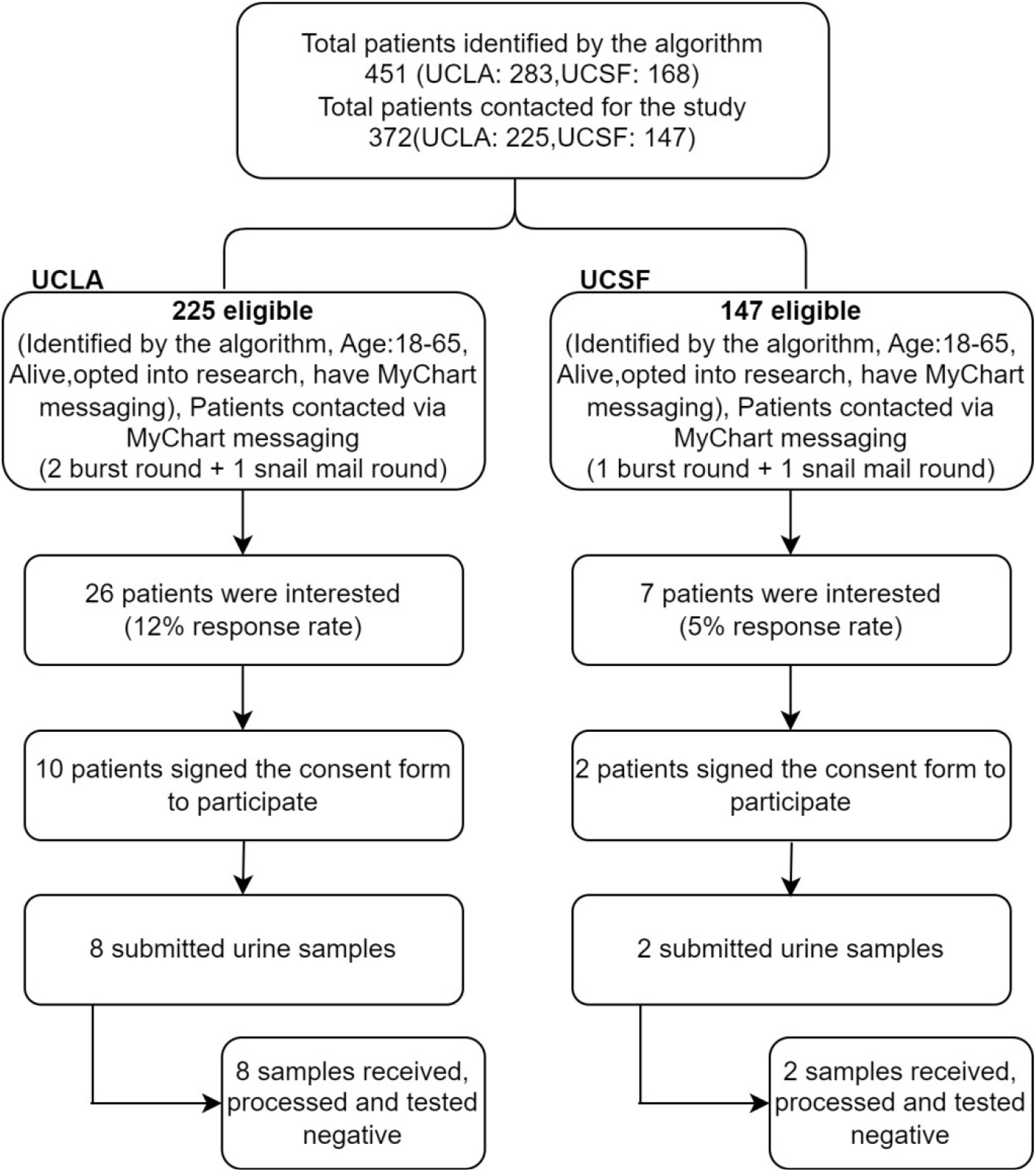
Summary of the prospective study and its outcomes. 451 patients were identified by the algorithm, of which 372 could be contacted for potential study enrollment. Enrollment was carried out in parallel in both participating medical centers. 10 subjects submitted specimens, all of which were negative.

### Model characterization

We assessed whether our models could have diagnosed any AHP case earlier than their true diagnosis date. We retrospectively applied our referral and diagnosis models to the available patient data as it existed at the end of each calendar year within the database, and calculated running probabilities of referral and diagnosis. We identified the year that a patient exceeded a referral probability of 10% and diagnosis probability of 50%, and compared it to their true diagnosis year by manual review (eFigure 3). We found that 71% of our cohort could have been diagnosed earlier than their true diagnosis year. The average reduction in the diagnostic delays of the total cohort was 1.2 years.

## DISCUSSION

We developed a new ML-based solution to reduce diagnostic delays in patients with rare diseases. Our strategy involved multiple steps. We used a causal model to identify gating steps along the patient journey and inform a modeling approach that avoids selection biases. We used cases of confirmed AHP from two academic medical centers to train models to sequentially predict patients who will be referred for a diagnostic testing as well as who will test positive, amongst those who are tested. Our models were highly accurate on internal validation, likely reflecting the use of automated ML and KG-enhanced feature engineering. Although our prospective validation effort was unsuccessful, it was to some extent expected for a rare disease. Moreover, it does not diminish the general conclusion that EHR data can play an important role in training algorithms to augment clinical decision making, enhance efficiencies within health systems, and improve patient outcomes. Indeed we found that 71% of our AHP cohort could be identified earlier than their actual diagnosis date, with a mean reduction of 1.2 years. Given the high burden of disease in symptomatic AHP patients and the availability of preventive treatment for acute attacks, such a reduction in diagnostic delays is likely to result in improved patient outcomes.

Our work contributes to the field of rare disease prediction by addressing weaknesses in the literature. The most direct precursor to our work is from Cohen et al^12^, who also developed an AHP prediction model using EHR data. Their method was limited by the lack of time-based preprocessing to prevent models from using “future” data elements (e.g. AHP treatments) to predict the “past” (e.g. an AHP diagnosis). This motivated our chosen approach. Many other similar efforts for other diseases have been published^13,22–26^, but none incorporate the full range of methods used here, including 1) notes-based features, 2) using domain knowledge and KGs to improve feature selection, 3) a two-stage modeling approach to avoid selection bias and model extrapolation, 4) using automated ML to optimize model performance, and 5) characterizing the model’s potential impact on the patient journey. This is the only rare disease prediction model we are aware of that has been subjected to a prospective test. Overall, we believe that this work sets a new, higher bar for the field of rare disease prediction and EHR-enabled research.

One limitation of our models is that they are unable to identify any patient with a near 0 probability of being diagnosed within a health system. This observation is related to the positivity assumption in causal inference^27^. Multiple social determinants of health may explain why a patient may never receive a within-system diagnosis: 1) the inability to afford the many visits needed to be recognized as a case (itself a potential consequence of unrecognized AHP), 2) concerns over being diagnosed with a hereditary and incurable condition, and 3) social circumstances that make patients less likely to consistently seek care at a single system. Other reasons could be provider related. AHP commonly presents in younger women, whose complaints may be written off and misdiagnosed. Although our models trained on a relatively diverse dataset, both are academic centers with a significant expertise, itself a gating factor in the patient journey (Figure 1).

Another limitation of this study was our inability to prospectively validate it. This reflected many factors, including 1) the rarity of AHP and thus the rarity of undiagnosed patients at UCSF/UCLA, and 2) institutional limitations against over-contacting patients for study recruitment. Future studies involving community systems and/or integrated networks with more longitudinal capture of the patient journey may address help these limitations and increase the chances of a successful validation.

In conclusion, we developed, characterized, and internally validated a novel machine learning approach to reduce diagnostic delays in AHP. Future work is needed to enhance these models over larger and more diverse datasets, prospectively validate its performance on undiagnosed patients, develop it into a decision support tool, and extend this approach to other diseases.

## Data Availability

The raw data used in this manuscript is available from UCSF and UCLA following the execution of a data use agreement. The analytical code is available at https://github.com/rwelab/AHPPrediction.

## Acknowledgements

The authors thank the contracting offices and Clinical and Translational Science Institutes of UCSF and UCLA for their support of this study. They thank the UCSF Academic Research Services and Bakar Computational Health Sciences Institute teams for supporting the computational needs of this study.

