## Supplemental data for "Reducing diagnostic delays in Acute Hepatic Porphyria using electronic health records data and machine learning: a multicenter development and validation study"

TABLE OF CONTENTS:

eFigure 1: Most predictive features in the diagnosis model

eFigure 2: Selection of the prospective cohort for model validation

eFigure 3: Running probabilities of a model-based diagnosis as compared to the actual diagnosis year.

eTable 1: Sample size calculations pertaining to a prospective test of the model accuracy

Supplemental Methods

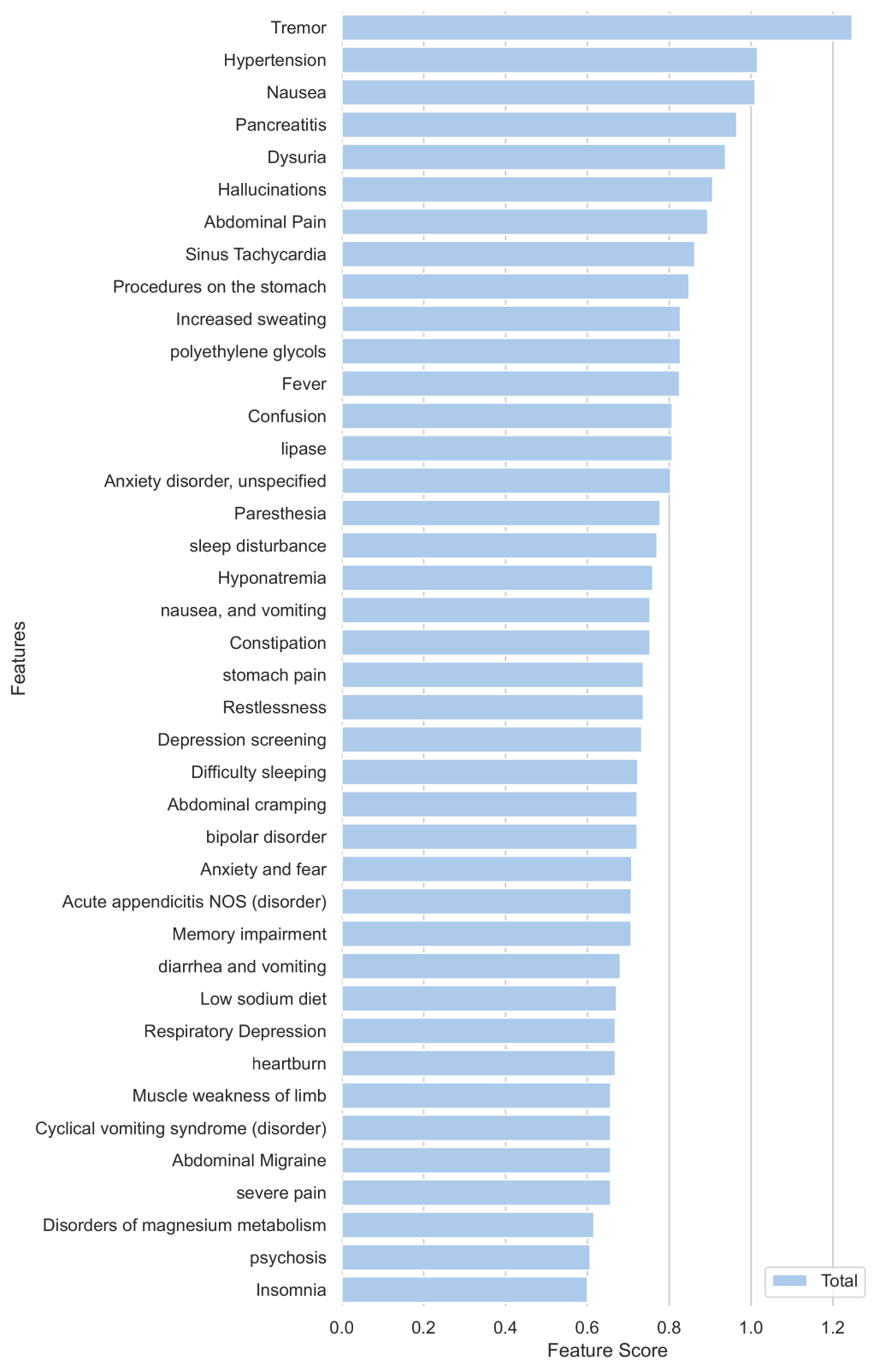

**eFigure 1**: Most predictive features in the diagnosis model

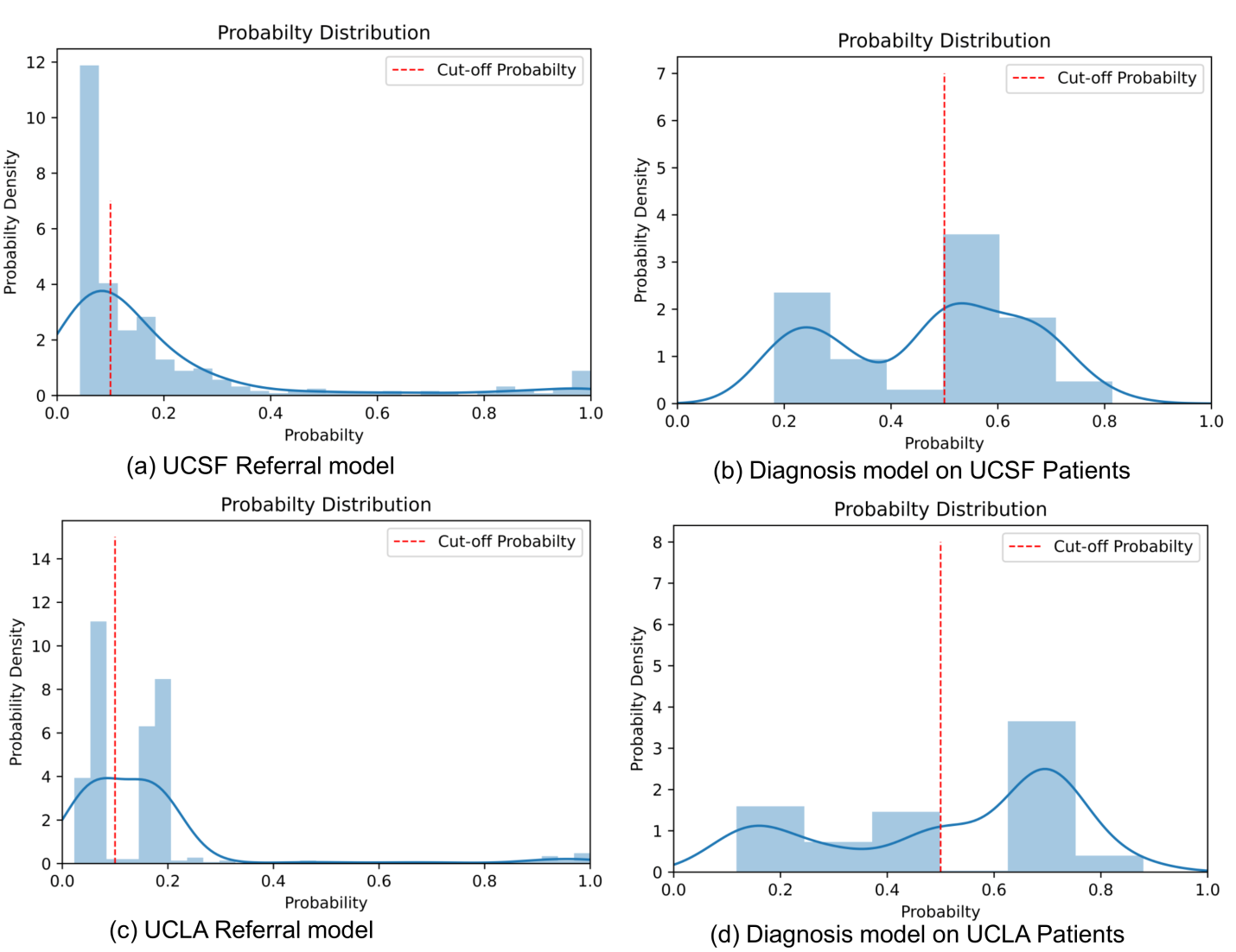

**eFigure 2**: Selection of the prospective cohort for model validation. From the pool of recently seen patients at UCSF and UCLA who lack an AHP diagnosis, we applied 10% referral probability and 50% diagnosis probability cutoffs (red lines) to identify potential recruitable subjects for a prospective test of our modeling framework.

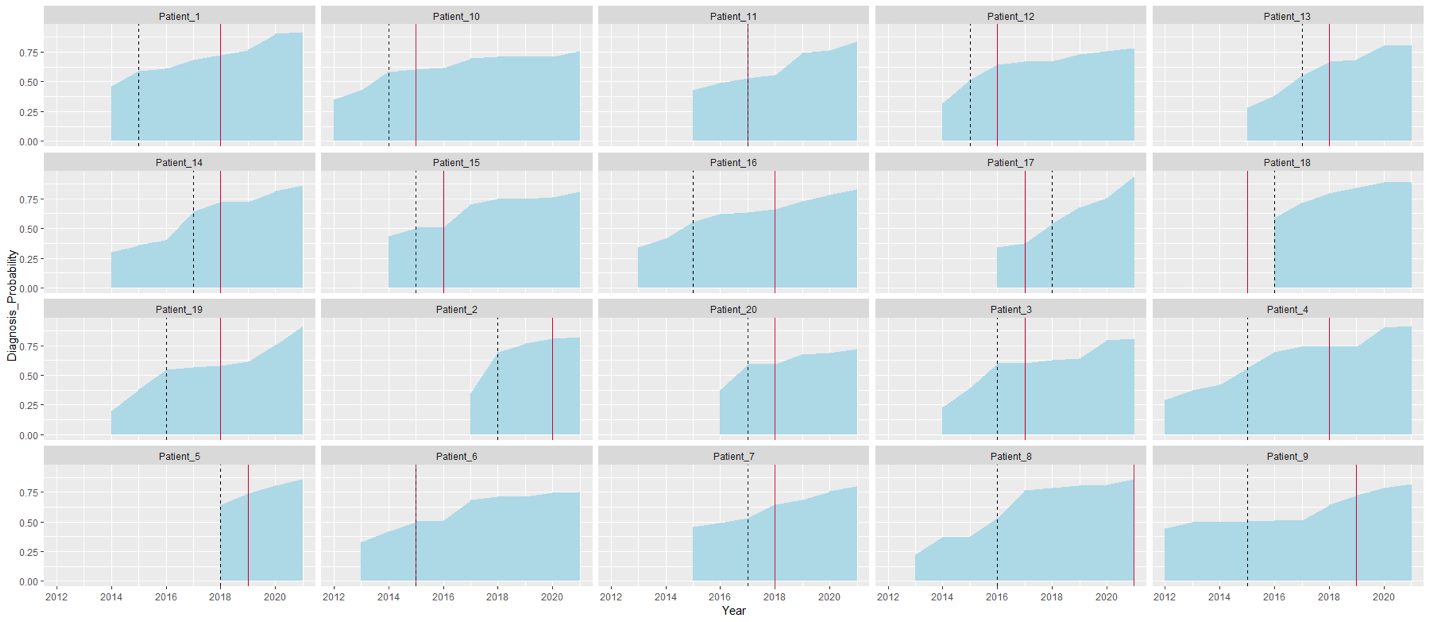

**eFigure 3**: Running probabilities of a model-based diagnosis as compared to the actual diagnosis year. Each plot represents the running probability of a model-based diagnosis in a different AHP patient. Because the diagnosis models are only suitable for use on a cohort of patients with a non-trivial probability of being tested for AHP (to limit extrapolation), all plots begin at the earliest year at which a given patient achieves a current referral probability ≥ 10%. The red solid line represents the actual year of diagnosis, whereas the black dotted line represents the earliest date that a patient is calculated as having a referral probability ≥ 10% and a diagnosis probability ≥ 50%. In cases where the black dotted line occurs earlier than the red solid line, the horizontal difference corresponds to potential years saved.

|  | High probability patient cohort | Expected number of new diagnoses | 95% confidence interval |
| --- | --- | --- | --- |
| UCSF | 168 | 31 | (22-42) |
| UCLA | 283 | 37 | (27-49) |
| Total | 451 | 68 | (54-84) |
| + Assuming 20% participation | NA | 14 | (7-24) |

**eTable 1**: Sample size calculations pertaining to a prospective test of the model accuracy

**Supplemental Methods:**

See our github page for the full code pipeline: <https://github.com/rwelab/AHPPrediction>. A few code snippets of interest to the feature engineering pipeline are reproduced below.

Python code snippet for processing EHR data from the 'Encounter' Table

df2.columns = ["PatientDurableKey", "PrimaryDiagnosisName", "PrimaryProcedureName","PrimaryProcedureCategory"]

df2['PrimaryDiagnosisName'] = df2['PrimaryDiagnosisName'].astype(str)+'_DX'

df2['PrimaryProcedureName'] = df2['PrimaryProcedureName'].astype(str)+'_PROC'

new = df2[['PatientDurableKey', 'PrimaryDiagnosisName']].copy()

new1 = df2[['PatientDurableKey', 'PrimaryProcedureName']].copy()

new.rename(columns={'PrimaryDiagnosisName': 'Feature'}, inplace=True)

new1.rename(columns={'PrimaryProcedureName': 'Feature'}, inplace=True)

dfq=new.groupby(["PatientDurableKey","Feature"]).size().reset_index(name="DepFreq")

dfw=new1.groupby(["PatientDurableKey","Feature"]).size().reset_index(name="DepFreq")

df_final=dfq.append(dfw, ignore_index=True)

df_final=df_final[~df_final.Feature.str.contains('\*Not Applicable')]

df_final=df_final[~df_final.Feature.str.contains('\*Unspecified')]

encounter=df_final

Code snippet for processing EHR from 'Medication' Table:

df5.columns = ["PatientDurableKey", "MedicationName", "MedicationTherapeuticClass", "MedicationPharmaceuticalClass" ]

new = df5[['PatientDurableKey', 'MedicationName']].copy()

new1 = df5[['PatientDurableKey', 'MedicationTherapeuticClass']].copy()

new2 = df5[['PatientDurableKey', 'MedicationPharmaceuticalClass']].copy()

new.rename(columns={'MedicationName': 'Feature'}, inplace=True)

new1.rename(columns={'MedicationTherapeuticClass': 'Feature'}, inplace=True)

new2.rename(columns={'MedicationPharmaceuticalClass': 'Feature'}, inplace=True)

dfq=new.groupby(["PatientDurableKey","Feature"]).size().reset_index(name="DepFreq")

dfw=new1.groupby(["PatientDurableKey","Feature"]).size().reset_index(name="DepFreq")

dfx=new2.groupby(["PatientDurableKey","Feature"]).size().reset_index(name="DepFreq")

df_final=dfq.append(dfw, ignore_index=True)

df_final=df_final.append(dfx, ignore_index=True)

df_final=df_final[~df_final.Feature.str.contains('\*Not Applicable')]

df_final=df_final[~df_final.Feature.str.contains('\*Unspecified')]

medication=df_final

medication.sort_values('DepFreq',ascending=False)

One hot encoding:

from sklearn.preprocessing import OneHotEncoder

encoder = OneHotEncoder(categories='auto')

\# fit the encoder to the data

encoder.fit(data)

\# transform the data

encoded_data = encoder.transform(data)
